# COVID-19 vaccine willingness and cannabis use histories

**DOI:** 10.1101/2021.04.26.21256109

**Authors:** Philip A. Spechler, Jennifer L. Stewart, Rayus Kuplicki, the Tulsa 1000 Investigators, Martin P. Paulus

## Abstract

**Background:** Cannabis use is associated with problematic health-behaviors such as excessive alcohol and tobacco use, and sedentary behavior. Here, we examined the association between cannabis use history and an especially topical health-behavior, willingness to receive a COVID-19 vaccine.

**Methods:** COVID-19 vaccine willingness was surveyed in a subset of participants from the Tulsa 1000 Study, which is a longitudinal study of psychiatric treatment-seeking and healthy control participants. We identified 45 participants who completed a COVID-19 vaccine questionnaire and reported more than 10 lifetime cannabis uses. Those participants were compared to a group of 45 individuals with very light (<10) cannabis use histories who were propensity score-matched on age, sex, income, and race. Two-group *t*-tests and Bayes factor analysis on vaccine willingness were conducted between groups. Exploratory correlation analyses were conducted on vaccine willingness and lifetime cannabis use levels within the cannabis group only.

**Results:** Vaccine willingness did not differ between the two groups (*t*_*88*_=0.33, *p*=.74; BF_01_=4.3). However, a negative correlation was identified within the cannabis group, such that higher lifetime cannabis use histories correlated with less willingness to receive a vaccine (rho_*43*_= -.33, *p*=.03).

**Conclusions:** Although vaccine willingness did not differ between the two matched groups, preliminary evidence suggests that heavy lifetime cannabis use might indicate a reluctance to engage in health-promoting behaviors like receiving a COVID-19 vaccine.

## 1.0 Introduction

On December 11^th^, 2020, the Food and Drug Administration issued an emergency use authorization for the Pfizer/BioNTech vaccine^1^, thus becoming the first vaccine to combat SARS-CoV-2 infections in the United States. One specific challenge facing vaccine uptake is the willingness of various subgroups of individuals to seek out and consent to vaccination^2^. A recent study from the Center for Disease Control (CDC) on intent to receive a COVID-19 vaccine found that vaccine willingness differed by sociodemographic factors. Generally, young adult women, non-Hispanic (black) individuals, and people from lower socioeconomic standings were less likely to express intent to become vaccinated^3^. Although the CDC did not stratify by drug use, a recent epidemiological study found that cannabis users comprise a similar sociodemographic profile^4^ to the CDC study on vaccine intent. Moreover, early data from Mellis and colleagues suggests that a quarter of people with substance use disorders were reportedly unwilling to receive a COVID-19 vaccine^5^. Hence, we hypothesized here that cannabis users may be reticent to receive the vaccine.

In light of early trends relating substance use with vaccine hesitancy^5^, substance use more generally might serve as an indicator variable reflecting a lower likelihood to engage in healthy behaviors. Previous research showed that relative to non-users, cannabis users were more likely to engage in excessive alcohol and tobacco use^6,7^, and have an unhealthy lifestyle like poor diet^8^, sedentary behavior^9^, and risky sexual and driving behaviors^10^. Similar individuals were also found to have lower rates of seeking preventative medical procedures like routine dental visits^11^, cancer screenings and influenza vaccinations^12,13^. Reasons for these findings are likely multifaceted, including the stigmatization of drug use^14^ and socioeconomic disparities that preclude access to healthcare. The present study seeks to understand if unwillingness to receive a COVID-19 vaccine might be one specific feature exhibited by cannabis users that reflects a lower propensity to engage in health-promoting behaviors.

Soon after the Pfizer/BioNTech COVID-19 vaccine was granted emergency use authorization, participants from our ongoing study of psychiatric treatment-seeking and healthy participants (the Tulsa 1000 Study^15^) were surveyed on their willingness to receive a COVID-19 vaccine. We then identified participants with cannabis use histories, and hypothesized that cannabis users would be less willing to receive a COVID-19 vaccine relative to non-using peers. The goal here is to understand if cannabis users are less likely to participate in a specific health-promoting behavior (i.e., vaccination). In doing so, we are careful to not dismiss or stigmatize individuals who are hesitant to receive a vaccine; individuals’ concerns related to a new vaccine should be compassionately addressed. The insights gained from this study might help guide targeted vaccine education efforts and encourage healthcare experts to have dialogues with these populations to increase vaccine uptake^16^. Overcoming issues of vaccine willingness is crucial in order to reach herd immunity levels in a timely fashion^17^, and to curb COVID-19 infection in vulnerable populations like substance users.

## 2.0 Methods

### 2.1 Data Collection

Participants from our longitudinal study of psychiatric populations (Tulsa 1000) were re-evaluated on their health-behaviors during the COVID-19 pandemic. Briefly, the Tulsa 1000 study is an ongoing project examining the biological and psychosocial features of treatment-seeking individuals with mood/anxiety, substance use, and eating disorders (plus healthy controls). See Victor et al., 2018 for an overview of the study^15^. All study procedures were approved by an institutional review board, and participants provided written informed consent.

### 2.2 Baseline Assessment

Lifetime cannabis use history was collected at baseline prior to the pandemic via the Customary Drinking and Drug Use Record (CDDR) survey^18^. Participants provided a quantitative free response, and those data were transformed onto a log10-scale due to departures from normality. Age, sex, income, and race were also collected via self-report at baseline, and the average time between baseline cannabis use and follow-up vaccine questionnaire measurement was four years and six months.

### 2.3 Follow-up Assessment

The follow-up assessment involved Remote data collection from December 28^th^, 2020 to January 27^th^, 2021 and was conducted using REDcap tools. Participants completed online questionnaires surveying their COVID-19 related experiences and health-behaviors, including their willingness to receive a COVID-19 vaccine. Specifically, participants were asked, “How much do you agree with the statement: I am willing to be vaccinated for COVID-19”. Responses were measured on an ordinal scale, from 1=completely disagree to 5=completely agree. A coarse measure of recent cannabis use was collected as part of the Coronavirus Health Impact Survey (CRISIS; http://www.crisissurvey.org). This survey asked how frequently the participant used cannabis in the past two weeks, and was measured on an ordinal scale, from 1=None to 5=Regularly. Current anxiety and depression symptom levels were also measured at follow-up using the Patient-Reported Outcomes Measurement Information System (PROMIS)^19^ questionnaires.

### 2.4 Selected Participants

Using data collected up to January 27^th^ 2021, a total of N=128 participants from the original T1000 sample had provided lifetime cannabis use data at baseline *and* responded to the COVID-19 vaccine questionnaire. From those 128, there were n=45 participants who had used cannabis more than 10 times in their lifetime, leaving n=82 participants with very light (≤10 lifetime uses) or no cannabis use histories to be selected from during matching. It was not possible to match on a group of cannabis-naïve participants only (zero lifetime uses), hence the threshold for lifetime uses was set at 10.

Next, a comparison group of individuals with very light cannabis use histories (n=45) was identified using propensity score-matching^20^ via the ‘MatchIt’ library in R. Variables used for matching included age, sex, income, and race (white vs. non-white). After running propensity score-matching, independent samples *t*-tests were used to confirm successful matching.

### 2.5 Statistical Analyses

Between-group comparisons on vaccine willingness was analyzed using an independent samples *t*-test. Bayes factor analysis was then used to quantify the level of support for the null (H_0_) vs. alternative (H_1_) hypothesis (hence, BF_01_) via the ‘BayesFactor’ library in R. Briefly, Bayes factors reflect the likelihood ratio between two competing hypotheses (null over alternative), with the general consensus being values >3 indicate ‘substantial’ support for the null^21^. Lastly, the correlation between vaccine willingness and lifetime cannabis use was calculated as an exploratory analysis within the cannabis use group only.

## 3.0 Results

Propensity score-matching successfully identified a similar group of individuals with very light cannabis use histories (n=45). Although this comparison group contained participants with some cannabis use (median 5 lifetime uses), the majority of individuals in this sample were cannabis-naïve (n=26, 58%). Examination of past two-week cannabis use data collected at the time of the vaccine survey confirmed that the cannabis group used more cannabis in the past two weeks relative to comparators (Mann-Whitney *U*=827, p=.04). However, most participants, regardless of group, reported no cannabis use in the past two weeks. Furthermore, although both groups contained individuals drawn from the mood/anxiety and healthy control populations from the larger T1000 study, participants from those populations were found to be balanced across the cannabis and comparison groups. See Table 1 for information on group characteristics and comparisons.

**Table 1:** Group characteristics

| Feature | Cannabis Users (N=45) |  |  |  |  | Matched Comparison Sample (N=45) |  |  |  |  | *p* |
| --- | --- | --- | --- | --- | --- | --- | --- | --- | --- | --- | --- |
| Age (years) (*M,SD*)<br>[Range] | 38.3, 11.1,<br>[18.6, 55.6] |  |  |  |  | 38.0, 11.4,<br>[18.3, 54.0] |  |  |  |  | .86 |
| Annual Income (*M,SD*)<br>[Range] | 58522, 44484,<br>[0,175000] |  |  |  |  | 61404, 49035,<br>[0,260000] |  |  |  |  | .77 |
| Sex (Male, Female) | 19, 26 |  |  |  |  | 18, 27 |  |  |  |  | .83 |
| Race (White, Non-White) | 34, 11 |  |  |  |  | 27, 18 |  |  |  |  | .18 |
| T1000 Group (Mood/Anxiety, Healthy Control) | 38, 7 |  |  |  |  | 34, 11 |  |  |  |  | .43 |
| Lifetime Cannabis Use (*M,SD*)<br>[Range] | 782.1, 1862.5,<br>[15, 9700] |  |  |  |  | 2.5, 3.5,<br>[0,10] |  |  |  |  | <.01 |
| Cannabis use in past two weeks | 1 | 2 | 3 | 4 | 5 | 1 | 2 | 3 | 4 | 5 | .04 |
| N | 31 | 5 | 1 | 2 | 6 | 39 | 3 | 0 | 1 | 2 |  |

Propensity score-matching procedure used age, income, sex, and race, as matching covariates to identify a similar sample of individuals with very light ( $\leq 10$ ) cannabis use histories. Lifetime cannabis use levels collected via the CDDR survey<sup>18</sup>. Cannabis use in past two weeks from CRISIS survey, where 1='None', 2='Rarely', 3='Occasionally', 4='Often', 5='Regularly', and was collected at the time of the vaccine willingness questionnaire *P*-values from two-group *t*-tests and chi-square tests (for sex, race) to confirm successful matching on first four variables.

Vaccine willingness did not significantly differ between the two matched groups (*t*_*88*_=.33, *p*=.74; Cannabis Group *M*=3.5, *SD*=1.6; Comparison Group *M*=3.6, *SD*=1.6). Bayes Factor analysis suggested these results were roughly four times more likely under the null hypothesis relative to the alternative (BF_01_=4.3). Lastly, within the cannabis group only, a significant correlation was identified between vaccine willingness and lifetime cannabis use levels (*r*_43_= -.33, *p*=.03; Figure 1). Despite the ordinal nature of the vaccine willingness data, results were reproduced using non-parametric statistics (Mann-Whitney *U*=985, *p*=.81; cannabis group rank correlation rho_43_=-.33, *p*=.03). Lastly, vaccine willingness was unrelated to current anxiety (*r*_43_=-.04, *p*=.77) or depression (*r*_43_=-.01, *p*=.95) levels within the cannabis group.

**Figure 1:**
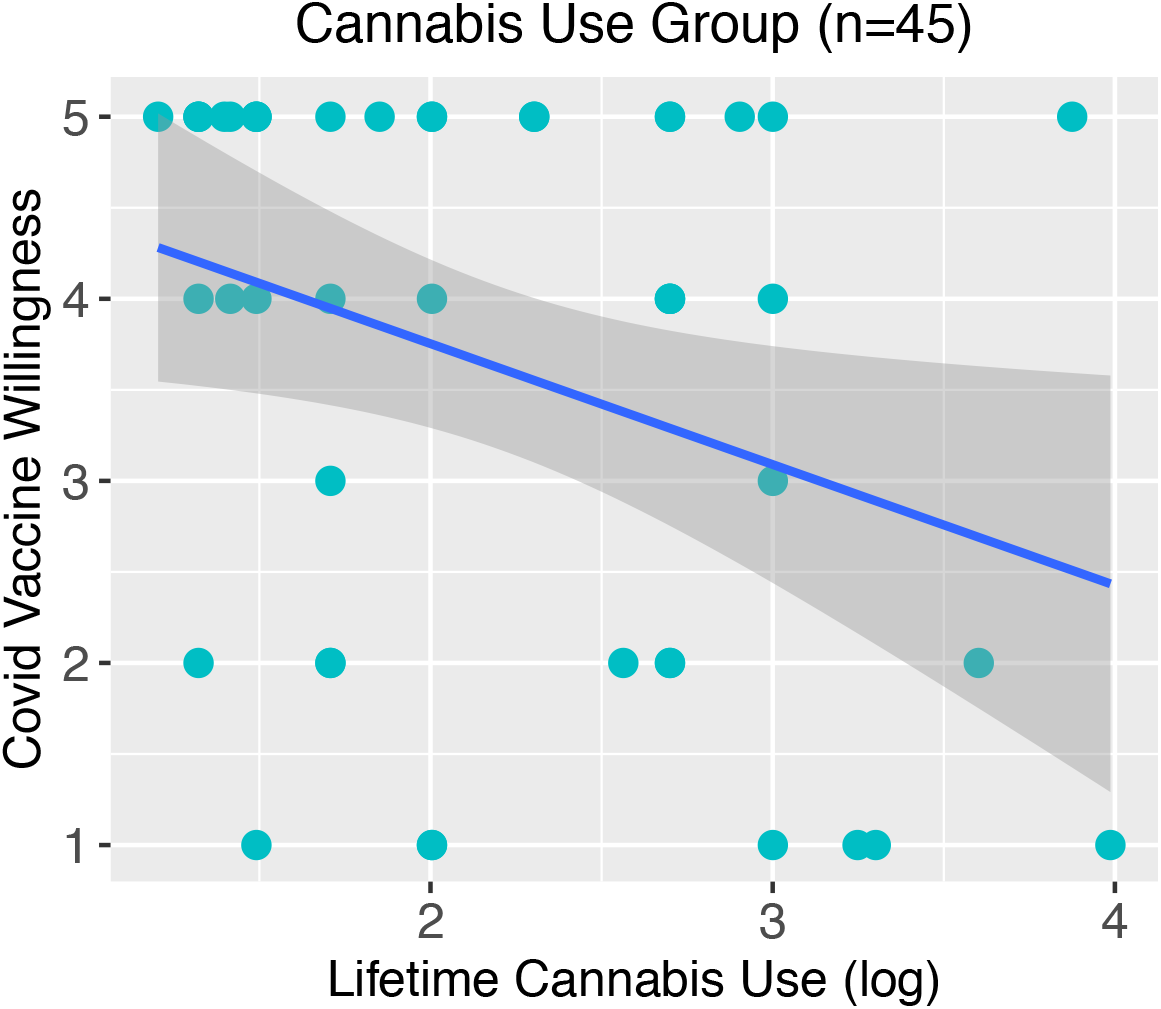
Correlation between vaccine willingness and lifetime cannabis use. Within the cannabis using group only, a negative correlation was identified (rho=-.33), suggesting that a higher cannabis use history was related to a lower likelihood of being willing to receive a COVID-19 vaccine.

## 4.0 Discussion

In this study, we hypothesized that a group of individuals with high cannabis use histories would be less willing to receive a COVID-19 vaccine relative to a similar group with very light cannabis use histories. Between-group analyses did not detect any differences on vaccine willingness when comparing the cannabis group to a sample matched on age, sex, income, and race. Bayes factor analysis provided substantial support for the null hypothesis. Despite the lack of a between-group difference, the mean values for vaccine willingness in each group suggested these participants were overall neutral-to-moderately willing to receive a vaccine.

Although the between-group results do not necessarily point to a specific effect of cannabis use, exploratory analyses restricted to the cannabis group suggested that higher lifetime cannabis use was associated with lower vaccine willingness. Therefore, heavy cannabis use might indicate a lower likelihood to engage in health-promoting behaviors. Coupled with the average vaccine hesitancy displayed by both groups, these results call for a need to encourage vaccine education and to address the concerns of these participants as a means to potentially increase vaccine willingness^16^. Efforts to readily vaccinate substance users are also needed because recent reviews suggest that COVID-19 might be more debilitating among these individuals due to psychosocial barriers to healthcare^22^ and an overrepresentation of comorbidities^23^ found among substance users.

Strengths of this study include the use of propensity score-matching, which made it unlikely that the matching variables confounded group membership or vaccine willingness^24^. Although the T1000 study included anxious/depressed and healthy control participants, the two groups studied here were found to be balanced on these populations (table 1), and vaccine willingness was unrelated to current anxiety and depression symptom levels. Nonetheless, it is possible that other psychiatric or physical health comorbidities unaccounted for may have influenced the results. An additional strength includes the use of Bayes factor analysis, which helped satisfy the shortcomings related to null-hypothesis significance testing^25^ and provided a more intuitive way to interpret the null findings.

Limitations of this study involve the heavy lifetime cannabis use levels within the cannabis group. Hence, it is unclear if similar effects would be found in people with intermediate cannabis use histories. Despite collecting a coarse measurement of recent cannabis use at the time of vaccine willingness questionnaire, a continuous measurement of cannabis use was only collected at baseline. Hence, this study is also limited by the time gap between measurement of those two features. Additionally, participants were not asked to elaborate on their reasons for or against becoming vaccinated, however, data collection occurred just as vaccines were becoming available and skepticism over a new vaccine may have been at its highest. Recent data from the CDC indicated that intent to become vaccinated against COVID-19 may increase with time^3^. Thus, it is possible that participants’ willingness may have evolved since data collection as they witnessed more people receiving a vaccine. Future studies are needed to continue to monitor COVID-19 vaccine intent and uptake, as well as infection rates, among substance users. These studies will help inform the epidemiological characteristics of COVID-19 within substance users, and provide insights into combatting the pandemic within vulnerable populations.

## Data Availability

The data and analysis codes that support the findings of this study are available from the corresponding author upon reasonable request.

## Acknowledgements

This work has been supported in part by The William K. Warren Foundation, and the National Institute of General Medical Sciences Center Grant Award Number 1P20GM121312. The content is solely the responsibility of the authors and does not necessarily represent the views of the WKW Foundation or the National Institutes of Health. The ClinicalTrials.gov identifier for the clinical protocol associated with data published in the current paper is NCT02450240, “Latent Structure of Multi-level Assessments and Predictors of Outcomes in Psychiatric Disorders.” The Tulsa 1000 Investigators include the following contributors: Robin Aupperle, Ph.D., Jerzy Bodurka, Ph.D., Savlador M. Guinjoan, M.D., Ph.D., Sahib S. Khalsa, M.D., Ph.D., Jonathan Savitz, Ph.D., Teresa A. Victor, Ph.D. This analysis was not pre-registered and the results should be considered exploratory.

